# Electronic Health Record-Based Prediction Models to Inform Decisions about HIV Pre-exposure Prophylaxis: A Systematic Review

**DOI:** 10.1101/2025.01.17.25320732

**Authors:** Afiba Manza-A. Agovi, Caitlin T. Thompson, Rachel J. Meadows, Yan Lu, Rohit P. Ojha

**Author notes:** Corresponding Author*: Manza Agovi, PhD, Center for Epidemiology & Healthcare Delivery Research JPS Health Network, 1500 South Main Street Fort Worth, TX 76104.

## Abstract

**Background:** Several clinical prediction models have been developed using electronic health records data to help inform decisions about HIV pre-exposure prophylaxis (PrEP) prescribing, but the characteristics and quality of these models have not been systematically assessed. We identified and critically appraised the characteristics and quality of studies reporting the development of electronic health records (EHR)-based models predicting HIV risk to inform decisions about PrEP prescribing.

**Methods:** We searched PubMed and the CINAHL databases between January 1, 2013 and June 19, 2023, with keywords related to EHR, HIV, and clinical prediction. We extracted data using the Critical Appraisal and Data Extraction for Systematic Reviews of Prediction Modelling Studies (CHARMS) checklist and assessed risk of bias using the Prediction model Risk Of Bias Assessment Tool (PROBAST) short form. We used narrative synthesis to describe characteristics and quality of eligible models.

**Results:** We identified 324 studies, of which 7 studies (resulting in 7 models) were eligible for our review. Several studies inadequately reported key components of the corresponding model. Most models were developed in the United States and used machine learning methods. The area under the receiver operating characteristic curve was reported for six models, which ranged between 0.77 and 0.89. All models had high risk of bias, primarily because of low events per variable and risk of overfitting.

**Conclusions:** We observed inadequate reporting of key components and high risk of bias across all EHR-based models. Future studies would benefit from following standard reporting guidelines and best practices for developing prediction models, which may strengthen the validity and applicability of EHR-based prediction models for informing decisions about HIV PrEP prescribing.

**Trial registration:** The review protocol was registered and published in PROSPERO (CRD42023428057)

## INTRODUCTION

HIV pre-exposure prophylaxis (PrEP) is recommended by the United States Preventive Services Task Force (USPSTF) for people at high risk of HIV infection [1], but PrEP prescribing is suboptimal [2]. A critical barrier to PrEP prescribing for healthcare providers is the identification of individuals who are likely to benefit from PrEP [3]. Consequently, the USPSTF identified the need to develop and validate decision support tools for identifying appropriate PrEP candidates [1].

Clinical prediction models are popular decision support tools, and several models have been developed to facilitate the identification of PrEP candidates. Nevertheless, some models require additional data collection and may not be acceptable to already burdened healthcare providers [4]. Clinical prediction models developed using routinely collected data circumvent the need for additional data collection and may be more readily adopted through integration in electronic health records (EHR) systems [5]. The characteristics, performance, and quality of currently available models developed using EHR data have not been systematically assessed. The findings from such an assessment may inform the prioritization of models for external validation, which is a critical step before implementation in practice. Therefore, we aimed to systematically identify and critically appraise the characteristics and quality of studies reporting the development of EHR-based models predicting HIV risk to inform decisions about PrEP prescribing.

## METHODS

This systematic review is reported according to the Preferred Reporting Items for Systematic Reviews and Meta-Analyses (PRISMA) Guidelines (see *Figure 1* and *Additional file 1*) [6]. For data extraction and critical appraisal, we followed the checklist for Critical Appraisal and Data Extraction for Systematic Reviews of Prediction Modelling Studies (CHARMS) [7]. The review protocol was registered and published in PROSPERO (CRD42023428057) [8].

**Figure 1.**
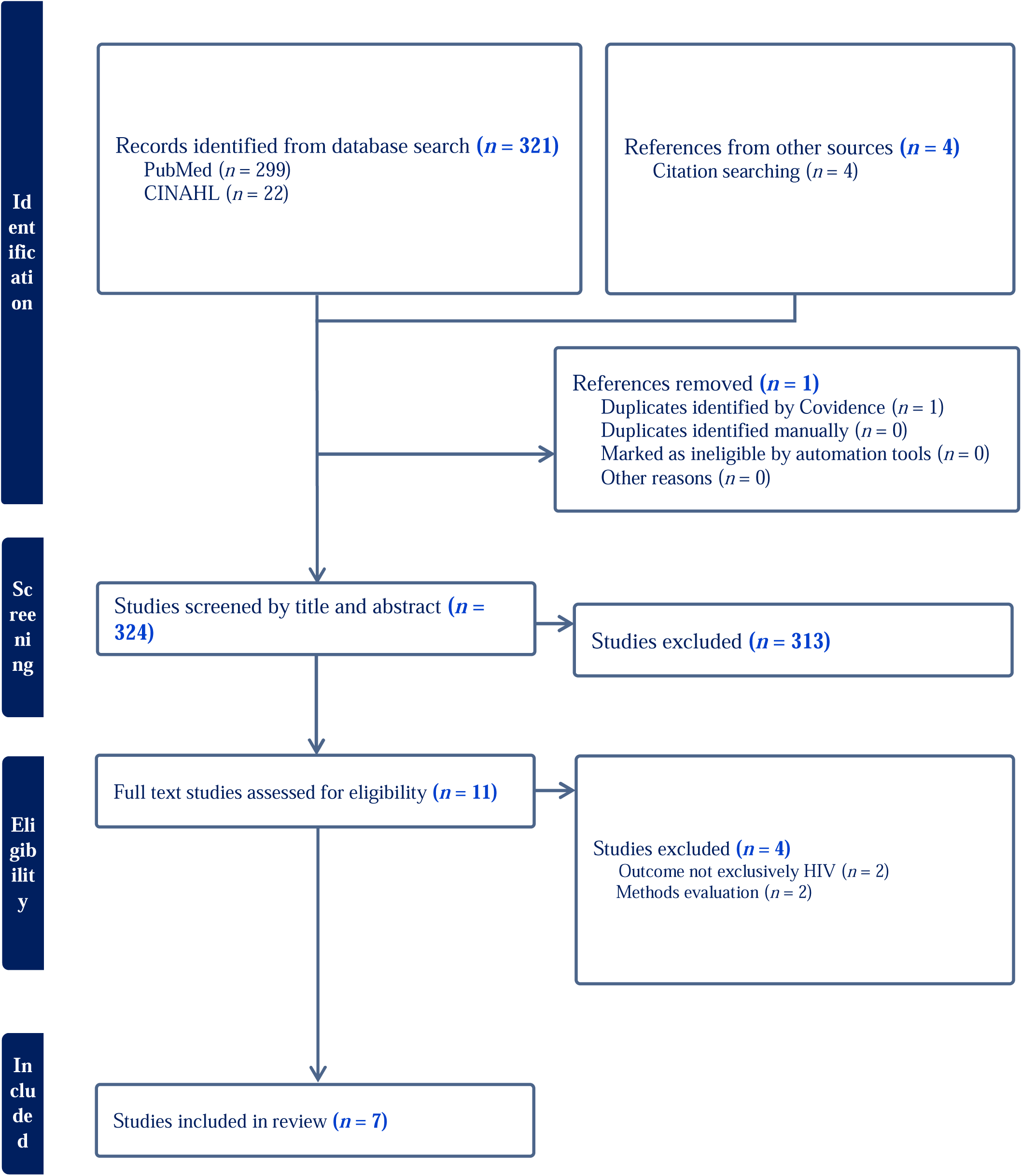
Preferred Reporting Items for Systematic Reviews and Meta-Analyses (PRISMA) flow diagram of study selection.

### Identification of prediction model studies

We systematically searched PubMed and the Cumulated Index to Nursing and Allied Health Literature (CINAHL) databases using a search strategy comprising keywords related to HIV and EHR (see *Appendix 1*). The key search terms in PubMed included the clinical queries broad filter for prediction models [9]. We developed our CINAHL search strategy using the same terms from our PubMed search. Our search included human studies that were published in English between January 1, 2013 and June 19, 2023 including Epub ahead of print articles. We also screened the reference lists of included studies to identify additional eligible citations not retrieved through the electronic search.

### Eligibility criteria

Articles were eligible for inclusion if: 1) published as an original research article; 2) based on any study design (e.g., cohort, case-control, etc.); 3) reported on the development or validation of a clinical prediction model using EHR data for identifying individuals aged ≥18 years with high risk of HIV infection and may benefit from PrEP; 4) the outcome of interest was exclusively HIV infection (*Appendix 2*). We excluded studies that did not exclusively use EHR data or were intended for methodologic evaluations rather than the development or validation of a potentially implementable model. In addition, we excluded review articles, editorials, letters to the editor, and conference abstracts. Lastly, if a study included variations of models, we only assessed the best performing model or the model recommended by study authors.

### Screening process

We used Covidence systematic review software [10] to screen and select eligible articles. Two reviewers (CTT and YL) independently screened article titles, abstracts, and full texts. Disagreements about article inclusion were resolved through discussion with two other investigators (AMA and RJM) before reaching a consensus.

### Data extraction

We extracted data for risk of bias assessment using a standardized data extraction form that was based on the CHARMS checklist [7], the Transparent Reporting of a multivariable prediction model for Individual Prognosis or Diagnosis (TRIPOD) checklist [11, 12], and the Prediction model Risk Of Bias Assessment Tool (PROBAST) [11]. The data extraction form included article publication information (author information, publication year, journal, country/location of publication), study characteristics (setting, source of data, sample characteristics), model development information (outcomes, candidate predictors, events per variable, sample size, missing data) and performance-related metrics (calibration, discrimination, classification measures, cut points for risk thresholds). We contacted authors for information or clarification if certain items were missing or unclear. Two reviewers (CTT and YL) independently performed the initial data extraction using our data extraction form. The extracted data were reviewed by two reviewers (AMA and RJM) to ensure accuracy, and discrepancies were resolved through a discussion with all four investigators (AMA, RJM, CTT, and YL).

### Risk of bias assessment

We used the PROBAST short form [11], which addresses two (outcome assessment and analysis) of the four key domains assessed in the full PROBAST [13], to assess the risk of bias for each article. The PROBAST short form is comparable to the full PROBAST for identifying prediction models with a high risk of bias [11]. The short form uses six questions from the full PROBAST that can be answered with “yes,” “no,” or “no information” (where “no” counts as 1). The scores from individual questions are summed for a total score of up to 6 points [11]. We described the overall risk of bias for each prediction model and each item in the two domains. We rated models with a total score of 0 points as low risk of bias, a total score of ≥1 as high risk of bias or high concerns, or unclear risk of bias if a model had “no information” on at least 1 item in a domain but none of the other domains were high risk of bias [11]. Four reviewers (AMA, RJM, CTT, and YL) independently performed the risk of bias assessment, and disagreements were resolved through discussion and consensus with the senior author (RPO).

### Evidence synthesis

We used narrative synthesis with descriptive statistics and data visualization to summarize information about eligible clinical prediction models. We did not pursue quantitative synthesis because our aim was to critically appraise eligible models. In addition, substantial heterogeneity between models precluded meaningful quantitative synthesis.

## RESULTS

### Study characteristics

Our search identified 324 de-duplicated records, of which seven articles (resulting in seven models) were included in our critical appraisal (*Figure 1)*. *Table 1* and *Table 2* summarize the characteristics of the seven included models, of which five (71%) were developed using data from the United States (US). Study settings included healthcare networks (n=5), a university hospital (n=1), and a public sexually transmitted infection (STI) clinic (n=1). Case-control was the most common study design (57%), and the remaining studies used a cohort design. Machine learning was the most common modelling method (n=5). Six models included internal validation and three models [14–16] also included external validation. Demographic information of the study populations was not consistently reported. For example, only four (57%) studies [14–17] reported participant age, where mean age ranged between 29 and 48 years. In addition, one study [18] reported a sex-specific prediction model which included only women. Three studies [15, 16] reported both internal and external validation, where external validation was based on temporal validation for two studies, and one study [14] performed both temporal and geographic validation.

**Table 1.** Characteristics of eligible studies reporting the development of electronic health record-based prediction models for HIV infection.

| Author (Year) | Study design | Study period | Study region | Participant characteristics | Female | Black | White | MSM<sup>c</sup> |
| --- | --- | --- | --- | --- | --- | --- | --- | --- |
|  |  |  |  | Age<sup>a</sup>, y |  |  |  |  |
| Burns et al. [17] (2022) | Cohort | 2014 – 2018 | US (Southeast) | 48 (23) | 56% | 27% | 63% | Not reported |
| Duthe et al. [19] (2021) | Case-control | 2013 – 2019 | Europe (West) | Not reported | 34% (cases) | Not reported | Not reported | 9% (cases) |
| Feller et al. [20] (2018) | Case-control | 2007 – 2015 | US (Northeast) | Not reported | 55% | Not reported | Not reported | Not reported |
| Friedman et al. [18] (2023) | Case-control | 2014 – 2020 | US (Midwest) | Not reported | 100% | 68% | 20% | Not applicable |
| Krakower et al. [14] (2019) | Case-control | 2007 – 2015 | US (Northeast) | 35 (22) | 57% | 5% | 60% | Not reported |
| Marcus et al. [15] (2019) | Cohort | 2007 - 2017 | US (West) | 45 (18) | 54% | 7% | 52% | Not reported |
| Xu et al. [16] (2022) | Cohort | 2015 – 2018 | Australia (Southeast) | 29 (8.1)<sup>b</sup> | 30% | Not reported | Not reported | 51% |

<sup>a</sup>Mean (standard deviation) or median (interquartile range)<sup>b</sup>Estimated from median and interquartile range[64]<sup>c</sup>MSM: Men who have sex with other men

**Table 2.** Characteristics of electronic health record-based prediction models for HIV infection.

| Author (Year) | Sample size | Events *n* (%) | Number of predictors |  | EPV<sup>b</sup> | Selection of final predictors | Number (%) of missing and missing data method | Type of validation | Performance measures<sup>a</sup> |
| --- | --- | --- | --- | --- | --- | --- | --- | --- | --- |
|  |  |  | Candidate | Final |  |  |  |  |  |
| Burns, 2022 | 998,787 | 117 (0.1) | 74 | 74 | 1.6 | Not reported | *n* (%): Not reported<br>Method: Missing indicator | Internal: Single random split-sample validation | *Discrimination*: AUC= 0.89 |
|  |  |  |  |  |  |  |  | External: None | None |
| Duthe, 2021 | 917 | 156 (17) | 162 | Not reported | 0.96 | LASSO | *n* (%): Not reported<br>Method: Not reported | Internal: Cross-validation | *Discrimination*: AUC=0.88, 95% CL: 0.81, 0.96 |
|  |  |  |  |  |  |  |  | External: None | None |
| Feller, 2018 | 724 | 181 (25) | 1,868 | 150 | 0.097 | Mutual information criteria | *n* (%): Not reported<br>Method: Not reported | Internal: Cross-validation | *Discrimination*: No conventional measure; F1 measure reported |
|  |  |  |  |  |  |  |  | External: None | None |
| Friedman, 2023 | 240 | 48 (20) | 24 | 12 | 2.0 | Univariable analysis | *n* (%): Not reported<br>Method: Complete-case analysis | Internal: None | *Discrimination*: AUC=0.74, 95% CL: 0.67, 0.81 |
|  |  |  |  |  |  |  |  | External: None | None |
| Marcus, 2019 | 3,143,963 | 701 (0.02) | 81 | 44 | 8.7 | LASSO | *n* (%): Not reported<br>Method: Added missing to reference category | Internal: Cross-validation | *Discrimination*: AUC=0.86, 95% CL: 0.85, 0.87 |
|  |  |  |  |  |  |  |  | External: Temporal | *Discrimination*: AUC=0.84, 95% CL: 0.80, 0.89<br>*Calibration*: Not evaluated |
| Xu, 2022 | 88,642 | 216 (0.2) | 35 | 10 | 6.2 | Variable importance analysis | *n* (%): Not reported<br>Method: Missing indicator | Internal: Cross-validation | *Discrimination*: AUC= 0.78, 95% CL: 0.72, 0.83 |
|  |  |  |  |  |  |  |  | External: Temporal (2019) | *Discrimination*: AUC= 0.79, 95% CL: 0.74, 0.84<br>*Calibration*: Not evaluated |
|  |  |  |  |  |  |  |  | External: Temporal (2020 – 2021) | *Discrimination*: AUC= 0.71, 95% CL: 0.60, 0.82<br>*Calibration*: Not evaluated |

**Table 2. Characteristics of electronic health record-based prediction models for HIV infection (continued).**
| Author<br>(Year) | Sample<br>size | Events<br>*n* (%) | Number of predictors |  | EPV<sup>a</sup> | Selection<br>of final<br>predictors | Number (%) of missing<br>and missing data method | Type of validation | Performance measures<sup>b</sup> |
| --- | --- | --- | --- | --- | --- | --- | --- | --- | --- |
|  |  |  | Candidate | Final |  |  |  |  |  |
| Krakower,<br>2019 | 7,616 | 150<br>(2.0) | 134 | 23 | 1.1 | LASSO | *n* (%): Not reported<br>Method: Not reported | Internal: Cross-validation | *Discrimination:* AUC=0.86,<br>95% CL: 0.82, 0.90 |
|  |  |  |  |  |  |  |  | External: Temporal | *Discrimination:* AUC=0.91,<br>95% CL: 0.81, 1.0<br>*Calibration:* 2.4/2.3 |
|  |  |  |  |  |  |  |  | External: Geographical | *Discrimination:* AUC=0.77,<br>95% CL: 0.74, 0.79<br>*Calibration:* Not evaluated |

<sup>a</sup>Performance measures correspond with the type of validation except for studies that did not undertake any validation. Apparent measures were reported for these studies.
<sup>b</sup>EPV: events per variable
<sup>c</sup>CL: confidence limits

### Outcome assessment

The outcome definition varied across models. Specifically, four models [14, 16, 17, 19] defined incident HIV as the initial positive HIV lab test, two models [15, 18] used local HIV registries for first lifetime HIV diagnosis, and one model [19] did not provide the criteria used to define incident HIV diagnosis. Only two models [15, 17] specified the timing of outcome measurement (within 1 to 3 years from baseline).

### Predictor selection and events per variable

Most models [14–17, 19, 20] reported selecting candidate predictors based on prior knowledge or literature review, but the approach to predictor selection was not reported for one model [18]. The most common predictors were age, sex, race/ethnicity, sexual behavior, substance use, and STIs (testing, diagnoses, and treatment). These data were extracted from EHR or clinical data warehouse. The number of candidate predictors, accounting for categorical predictors, in model development ranged between 24 and 1,868. Predictor selection was based on least absolute shrinkage and selection operator (LASSO) for three models [14, 15, 19], variable importance analysis for three models [16, 17, 20], and bivariable screening for one model [18]. The number of predictors included in the final models ranged from 10 to 150, but one model [19] did not report the number of final predictors. All models were based on less than 10 events per variable (EPV).

### Continuous predictors

Continuous predictors were categorized in 71% of the studies [15–18, 20], but no information was provided for any model about how cut points for categories were selected. Continuous predictors were not categorized in one study [14], and one study [19] provided no information on how continuous predictors were handled.

### Missing values

None of the studies reported the number of participants with missing values for predictors, but four models [15–18] discussed how missing data were handled. The missing indicator method was used for three models [15–17] and complete case analysis was used for one model [18].

### Overoptimism/overfitting

Studies reported internal validation of models using primarily cross-validation (five models) [14–16, 19, 20] One model used a random split sample [17], whereas another model did not use any method for internal validation [18]. Nevertheless, only four models [14–16, 19] used appropriate methods for adjusting overfitting and overoptimism.

### Overall risk of bias

*Figure 2* illustrates the risk of bias in EHR-based prediction models for HIV infection (see *Appendix 4* for details about model-specific biases). All prediction models had a high risk of bias overall and specifically in the outcome and analysis domains. Studies were rated as having a high risk of bias because of low EPV (highest EPV was 8.7), and all models either handled missing data inappropriately [15, 16, 18], or did not explicitly mention the approach to missing data [14, 19, 20]. In the outcome assessment domain, one study [18] provided no information on how the outcome was assessed. One study [17] included participants who had already been prescribed PrEP.

**Figure 2.**
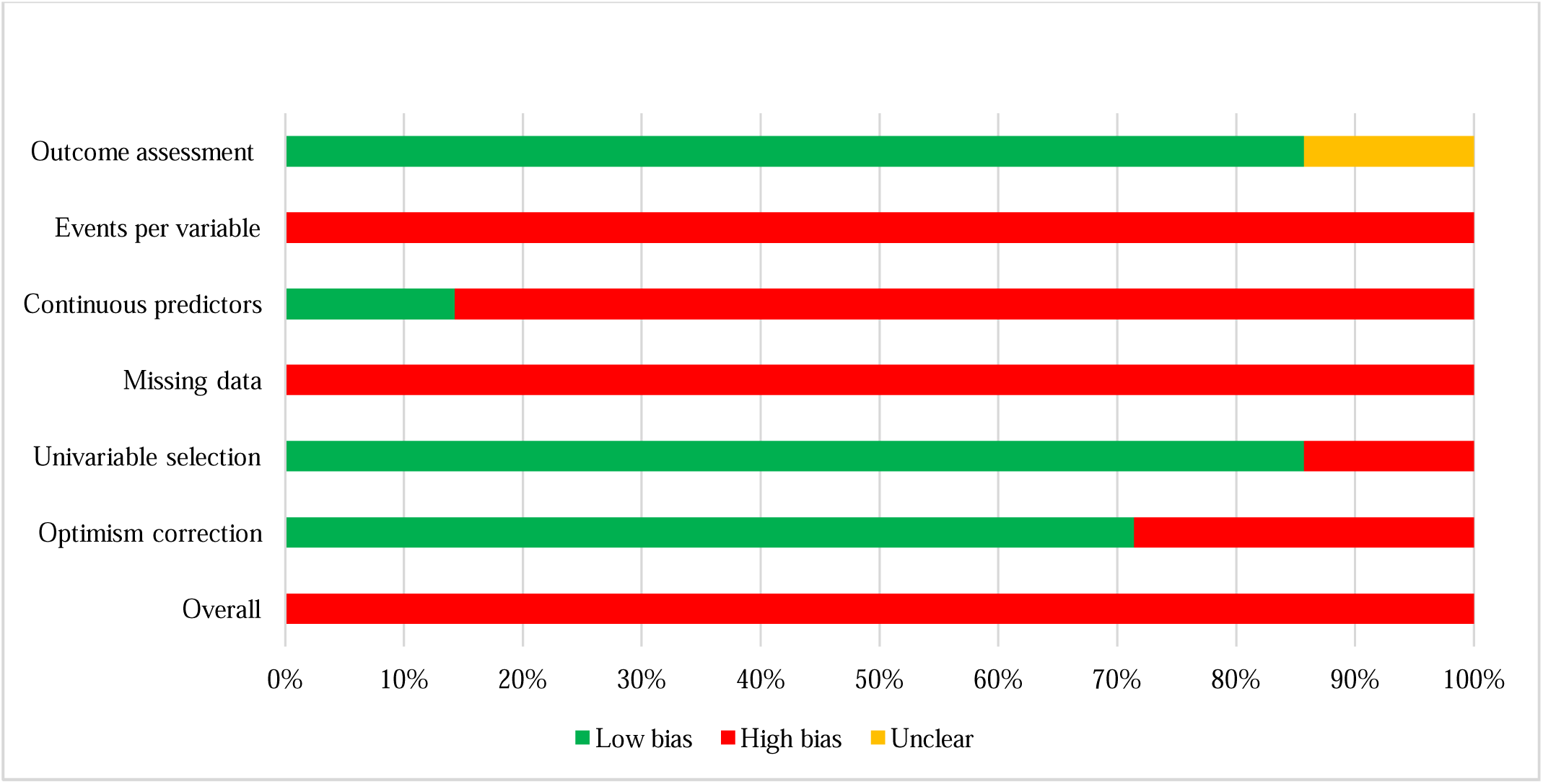
Prediction model Risk Of Bias Assessment Tool (PROBAST) short form results on risk of bias in identified electronic health record-based models for predicting HIV risk. The short form evaluates six items—outcome assessment, events per variable (EPV), continuous predictors, missing data, univariable selection and correction for overfitting/optimism—in the outcome and analysis domains. Color-coded bars show the proportion of studies at low (green), unclear (yellow), and high (red) risk of bias.

### Model performance

Six studies assessed discrimination based on the area under the receiver operating characteristic curve (AUC), whereas one study [20] assessed discrimination based on an F1-measure from a precision-recall curve. AUC values from internal validation ranged from 0.77 [14] to 0.89 [17] across studies. Only one study [14] that reported external validation also reported calibration. Krakower et al. [14] reported calibration-in-the-large (i.e., model-based predicted probability compared with observed probability of HIV incidence for the overall population).

## DISCUSSION

We identified and critically appraised seven clinical prediction models developed using EHR data that aimed to predict HIV risk and inform decisions about HIV PrEP prescribing. Most models were developed using machine learning methods and in US populations. Our critical appraisal indicates a high risk of bias in all reviewed models. In addition, few models have been evaluated for external validity.

### Limitations

Our findings should be interpreted in the context of certain limitations. Our search strategy focused on PubMed and CINAHL. We could have missed eligible models that were indexed outside of these literature databases. In addition, we only included studies that were published in English.

Nevertheless, searching two literature databases is generally sufficient, [21, 22] and recent evidence suggests that English language restrictions do not introduce consequential bias [23, 24]. Another consideration is that we restricted our search to models published after PrEP was approved in 2012 [25], which could overlook models published before 2013. We speculate that this restriction may have limited consequences, considering that most studies we identified were conducted in the U.S., where clinical guidelines for PrEP were published in 2014 [11] and large-scale transitions to EHR systems did not occur until 2015 [26].

### Evidence synthesis

The clinical prediction models included in our review presented challenges with appraisal because of inadequate reporting, which is pervasive across clinical topics regardless of whether the models were developed using statistical or machine learning methods [27–31]. Only one study reported adequate information for all components related to the risk of bias assessment using the PROBAST short form. For example, one report did not provide sufficient information about how outcomes were assessed, and most reports did not provide adequate information about model specification, including how continuous predictors were handled in the model. In addition, standard measures for model discrimination were not reported in one study, which reflects broader trends in inadequate reporting of models developed using machine learning methods [27]. The AUC is a standard measure of discrimination and should be reported over measures such as the area under the precision-recall curve (AUPRC), which has limited empirical justification and may exacerbate algorithmic biases [32]. Ultimately, inadequate reporting of prediction models hinders assessment, reproducibility, and implementation in clinical practice. Inadequate reporting for some study components also contributes to high risk of bias [11].

Our critical appraisal suggests high risk of bias in the development of all EHR-based models for predicting HIV infection included in our review. A key limitation across all models was overfitting given that the highest EPV for any model was 8.7, which indicates that the models included too many predictors relative to the observed number of incident HIV events. Overfitted models have a high risk of overoptimism (i.e., overestimated model performance) in external populations. The EPV threshold to avoid overfitting will vary across models based on factors such as sample size, predictor effects, predictor transformations, and outcome frequency [33–36], which emphasizes the need for appropriate estimation and adjustment for overoptimism [37, 38]. Nevertheless, models developed using conventional statistical methods typically stabilize when EPV exceeds 20, whereas models based on machine learning methods typically stabilize when EPV exceeds 200 [39]. None of the reviewed models approached these corresponding thresholds. Several models applied penalized methods (e.g., LASSO) to reduce overoptimism, but these methods are not necessarily effective, particularly when EPV is low [40, 41]. In addition, almost all models categorized continuous predictors. Categorization of continuous predictors is often arbitrary, assumes homogeneity within categories, and results in loss of information, power, and efficiency [42–45]. Several alternatives to categorization have been described in detail elsewhere [42, 43, 46].

The models identified in our review also present challenges with applicability. For example, no reports explicitly stated the intended moment of use of the model (i.e., prediction baseline), which creates uncertainty about when to measure predictors and use the model for clinical decision-making [47]. Specifying the moment of intended use is particularly important for reducing the potential for temporal bias, where predictors may be measured at some post-baseline time when clinicians would not have access to the information and predictors measured at a time closer to outcome occurrence will have stronger relations with the outcome [47, 48]. In addition, few models specified the prediction horizon (i.e., follow-up period for outcome occurrence such as 1-year risk of HIV), which creates ambiguity about the period relevant to model predictions. Lastly, the large number of predictors in several models may create challenges for applicability if some predictors are unmeasured or inconsistently measured in certain healthcare settings. Models that use predictors commonly and consistently measured across healthcare settings may be more widely applicable.

Our critical appraisal focused on the development of HIV risk prediction models, but external validation is the next critical step before implementing in practice [49–52]. External validation was conducted for three models [14–16] identified in our review. External validation involved temporal (i.e., same setting but different time period) and geographical validation (i.e., different geographic location) for one model [14] and temporal validation for the other models [15, 16]. Nevertheless, the outcome frequencies were too small for reliable external validation in both studies. For example, at least 100 events are needed for reliable external validation [53], but more precise computations are available for specifying minimum sample sizes and events [54, 55]. In addition, discrimination may be the primary focus during model development, but calibration is arguably more important during external validation because of the implications for clinical decision-making [56–58]. All three reports of models with external validation mentioned assessing calibration, but specific calibration measures were reported for only one model [14, 59].

## CONCLUSION

We systematically identified and appraised EHR-based clinical prediction models for HIV infection to inform decisions about HIV PrEP prescribing in healthcare settings. We observed inadequate reporting of key components across models. Reporting quality may be improved by adhering to the TRIPOD+AI guidelines [60] for models developed using machine learning or conventional statistical methods. Our critical appraisal suggested high risk of bias for all models. One critical source of bias was low EPV, which results in overfitted models and increases the risk of overoptimism [33–36, 38]. One approach is to identify a parsimonious core set of predictors relevant to incident HIV infection that are consistently measured across diverse healthcare settings, which would reduce the potential for overfitting and improve applicability [38]. In addition, data-sharing and open collaboration can facilitate aggregation of datasets for larger sample sizes and event numbers to increase EPV. Future studies should also refer to the PROBAST [13] or PROBAST-AI tool [61] currently under development to avoid common sources of bias when developing and validating prediction models. Lastly, models should specify the intended moment of use and prediction horizon to improve applicability.

EHR-based HIV prediction models could be useful tools for decisions about PrEP prescribing, but the consequences of improperly developed HIV prediction models are not trivial. Prediction models that underestimate HIV risk could result in missed prevention opportunities and leave individuals vulnerable to HIV infection. Conversely, overestimating HIV risk could result in unnecessary PrEP prescriptions, potential side effects, and financial burden from costly prescriptions [62]. Out-of-pocket costs for PrEP prescriptions are particularly concerning in states challenging no patient cost-sharing mandates for PrEP [63]. Consequently, proper development and validation of EHR-based HIV prediction models is necessary to ensure appropriate prescribing.

## ABBREVIATIONS

AUC: area under the receiver operating characteristic curve; AUPRC: Area under the precision-recall curve; AI: Artificial intelligence; CINAHL: Cumulated Index to Nursing and Allied Health Literature CHARMS: Critical Appraisal and Data Extraction for Systematic Reviews of Prediction Modelling Studies; EHR: Electronic Health Records; EPV: events per variable; PROBAST: Prediction model Risk Of Bias Assessment Tool; PrEP: Pre-exposure prophylaxis; TRIPOD: Transparent Reporting of a multivariable prediction model for Individual Prognosis or Diagnosis; PRISMA: Preferred Reporting Items for Systematic Reviews and Meta-Analyses.

## DECLARATIONS

### Ethics approval and consent to participate

Not applicable

### Consent for publication

Not applicable

### Availability of data and materials

The data that support the findings of this study are publicly available.

### Competing interests

The authors declare no conflict of interest.

### Funding

This research did not receive any specific grant from funding agencies in the public, commercial, or not-for-profit sectors.

## Author contributions

All authors contributed to the interpretation of the findings, critically revised the work for intellectual content, and approved the final version submitted for publication. AMA: Conceptualization, methodology, project administration, supervision, formal analysis, validation, writing-original draft, writing-review and editing. RPO: Conceptualization, methodology, writing-review and editing, supervision. RJM: Formal analysis, validation, writing-original draft, writing-review and editing. supervision. CTT: Data curation, formal analysis, visualization, writing-original draft, writing-review and editing. YL: Data curation, formal analysis, visualization, writing-original draft, writing-review and editing.

## Supporting information

PRISMA CHECKLIST

## Data Availability

The data that support the findings of this study are publicly available.

## Acknowledgements

The authors are grateful to Meng Pan, who contributed to protocol development.

## Notes

### Competing Interest Statement

The authors have declared no competing interest.

## REFERENCES

1. Owens DK, Davidson KW, Krist AH, Barry MJ, Cabana M, Caughey AB, et al. Preexposure Prophylaxis for the Prevention of HIV Infection: US Preventive Services Task Force Recommendation Statement. JAMA. 2019;321(22):2203–13.

2. Agovi AM, Anikpo I, Cvitanovich MJ, Fasanmi EO, Ojha RP, Marcus JL. Brief Report: HIV Pre-exposure Prophylaxis Prescribing in an Urban Safety-Net Health System. J Acquir Immune Defic Syndr. 2021;88(3):e17–e21.

3. Agovi AM, Anikpo I, Cvitanovich MJ, Craten KJ, Asuelime EO, Ojha RP. Knowledge needs for implementing HIV pre-exposure prophylaxis among primary care providers in a safety-net health system. Prev Med Rep. 2020;20:101266.

4. Kennedy G, Gallego B. Clinical prediction rules: A systematic review of healthcare provider opinions and preferences. Int J Med Inform. 2019;123:1–10.

5. Sharma V, Ali I, van der Veer S, Martin G, Ainsworth J, Augustine T. Adoption of clinical risk prediction tools is limited by a lack of integration with electronic health records. BMJ Health Care Inform. 2021;28(1).

6. Moher D, Liberati A, Tetzlaff J, Altman DG, Group P. Preferred reporting items for systematic reviews and meta-analyses: the PRISMA statement. BMJ. 2009;339:b2535.

7. Moons KG, de Groot JA, Bouwmeester W, Vergouwe Y, Mallett S, Altman DG, et al. Critical appraisal and data extraction for systematic reviews of prediction modelling studies: the CHARMS checklist. PLoS Med. 2014;11(10):e1001744.

8. Agovi A, Thompson CT, Pan M, Ojha RP. Electronic Health Record-based Prediction Models to Identify Potential Candidates for HIV Pre-exposure Prophylaxis: A Systematic Review. 2023.

9. Wong SS, Wilczynski NL, Haynes RB, Ramkissoonsingh R, Hedges T. Developing optimal search strategies for detecting sound clinical prediction studies in MEDLINE. AMIA Annu Symp Proc. 2003;2003:728–32.

10. Covidence systematic review software. Melbourne, Australia.: Veritas Health Innovation.

11. Venema E, Wessler BS, Paulus JK, Salah R, Raman G, Leung LY, et al. Large-scale validation of the prediction model risk of bias assessment Tool (PROBAST) using a short form: high risk of bias models show poorer discrimination. J Clin Epidemiol. 2021;138:32–9.

12. Collins GS, Reitsma JB, Altman DG, Moons KG. Transparent reporting of a multivariable prediction model for individual prognosis or diagnosis (TRIPOD): the TRIPOD statement. BMJ. 2015;350:g7594.

13. Wolff RF, Moons KGM, Riley RD, Whiting PF, Westwood M, Collins GS, et al. PROBAST: A Tool to Assess the Risk of Bias and Applicability of Prediction Model Studies. Ann Intern Med. 2019;170(1):51–8.

14. Krakower DS, Gruber S, Hsu K, Menchaca JT, Maro JC, Kruskal BA, et al. Development and validation of an automated HIV prediction algorithm to identify candidates for pre-exposure prophylaxis: a modelling study. Lancet HIV. 2019;6(10):e696–e704.

15. Marcus JL, Hurley LB, Krakower DS, Alexeeff S, Silverberg MJ, Volk JE. Use of electronic health record data and machine learning to identify candidates for HIV pre-exposure prophylaxis: a modelling study. Lancet HIV. 2019;6(10):e688–e95.

16. Xu X, Yu Z, Ge Z, Chow EPF, Bao Y, Ong JJ, et al. Web-Based Risk Prediction Tool for an Individual’s Risk of HIV and Sexually Transmitted Infections Using Machine Learning Algorithms: Development and External Validation Study. J Med Internet Res. 2022;24(8):e37850.

17. Burns CM, Pung L, Witt D, Gao M, Sendak M, Balu S, et al. Development of a Human Immunodeficiency Virus Risk Prediction Model Using Electronic Health Record Data From an Academic Health System in the Southern United States. Clin Infect Dis. 2023;76(2):299–306.

18. Friedman EE, Shankaran S, Devlin SA, Kishen EB, Mason JA, Sha BE, et al. Development of a predictive model for identifying women vulnerable to HIV in Chicago. BMC Womens Health. 2023;23(1):313.

19. Duthe JC, Bouzille G, Sylvestre E, Chazard E, Arvieux C, Cuggia M. How to Identify Potential Candidates for HIV Pre-Exposure Prophylaxis: An AI Algorithm Reusing Real-World Hospital Data. Stud Health Technol Inform. 2021;281:714–8.

20. Feller DJ, Zucker J, Yin MT, Gordon P, Elhadad N. Using Clinical Notes and Natural Language Processing for Automated HIV Risk Assessment. J Acquir Immune Defic Syndr. 2018;77(2):160–6.

21. Ewald H, Klerings I, Wagner G, Heise TL, Stratil JM, Lhachimi SK, et al. Searching two or more databases decreased the risk of missing relevant studies: a metaresearch study. J Clin Epidemiol. 2022;149:154–64.

22. Halladay CW, Trikalinos TA, Schmid IT, Schmid CH, Dahabreh IJ. Using data sources beyond PubMed has a modest impact on the results of systematic reviews of therapeutic interventions. J Clin Epidemiol. 2015;68(9):1076–84.

23. Nussbaumer-Streit B, Klerings I, Dobrescu AI, Persad E, Stevens A, Garritty C, et al. Excluding non-English publications from evidence-syntheses did not change conclusions: a meta-epidemiological study. J Clin Epidemiol. 2020;118:42–54.

24. Morrison A, Polisena J, Husereau D, Moulton K, Clark M, Fiander M, et al. The effect of English-language restriction on systematic review-based meta-analyses: a systematic review of empirical studies. Int J Technol Assess Health Care. 2012;28(2):138–44.

25. U.S. Food and Drug Administration. FDA approves first drug for reducing the risk of sexually acquired HIV infection 2012 [Available from: https://web.archive.org/web/20121119085625/ http://www.fda.gov/NewsEvents/Newsroom/PressAnnouncements/ucm312210.htm.

26. Centers for Medicare and Medicaid Services. Medicare and Medicaid programs; electronic health record incentive program. Final rule. Fed Regist. 2010;75(144):44313–588.

27. Andaur Navarro CL, Damen JAA, Takada T, Nijman SWJ, Dhiman P, Ma J, et al. Completeness of reporting of clinical prediction models developed using supervised machine learning: a systematic review. BMC Med Res Methodol. 2022;22(1):12.

28. Yang C, Kors JA, Ioannou S, John LH, Markus AF, Rekkas A, et al. Trends in the conduct and reporting of clinical prediction model development and validation: a systematic review. J Am Med Inform Assoc. 2022;29(5):983–9.

29. Heus P, Damen J, Pajouheshnia R, Scholten R, Reitsma JB, Collins GS, et al. Poor reporting of multivariable prediction model studies: towards a targeted implementation strategy of the TRIPOD statement. BMC Med. 2018;16(1):120.

30. Zamanipoor Najafabadi AH, Ramspek CL, Dekker FW, Heus P, Hooft L, Moons KGM, et al. TRIPOD statement: a preliminary pre-post analysis of reporting and methods of prediction models. BMJ Open. 2020;10(9):e041537.

31. Bouwmeester W, Zuithoff NP, Mallett S, Geerlings MI, Vergouwe Y, Steyerberg EW, et al. Reporting and methods in clinical prediction research: a systematic review. PLoS Med. 2012;9(5):1–12.

32. McDermott M, Hansen LH, Zhang H, Angelotti G, Gallifant J. A Closer Look at AUROC and AUPRC under Class Imbalance. arXiv preprint arXiv:240106091. 2024.

33. Riley RD, Ensor J, Snell KIE, Harrell FE, Jr., Martin GP, Reitsma JB, et al. Calculating the sample size required for developing a clinical prediction model. BMJ. 2020;368:m441.

34. Riley RD, Snell KIE, Ensor J, Burke DL, Harrell FE, Jr., Moons KGM, et al. Minimum sample size for developing a multivariable prediction model: Part I - Continuous outcomes. Stat Med. 2019;38(7):1262–75.

35. Riley RD, Snell KI, Ensor J, Burke DL, Harrell FE, Jr., Moons KG, et al. Minimum sample size for developing a multivariable prediction model: PART II - binary and time-to-event outcomes. Stat Med. 2019;38(7):1276–96.

36. van Smeden M, de Groot JA, Moons KG, Collins GS, Altman DG, Eijkemans MJ, et al. No rationale for 1 variable per 10 events criterion for binary logistic regression analysis. BMC Med Res Methodol. 2016;16(1):163.

37. Austin PC, Steyerberg EW. Events per variable (EPV) and the relative performance of different strategies for estimating the out-of-sample validity of logistic regression models. Stat Methods Med Res. 2017;26(2):796–808.

38. Steyerberg EW, Uno H, Ioannidis JPA, van Calster B, Collaborators. Poor performance of clinical prediction models: the harm of commonly applied methods. J Clin Epidemiol. 2018;98:133–43.

39. van der Ploeg T, Austin PC, Steyerberg EW. Modern modelling techniques are data hungry: a simulation study for predicting dichotomous endpoints. BMC Med Res Methodol. 2014;14:137.

40. Riley RD, Snell KIE, Martin GP, Whittle R, Archer L, Sperrin M, et al. Penalization and shrinkage methods produced unreliable clinical prediction models especially when sample size was small. J Clin Epidemiol. 2021;132:88–96.

41. Van Calster B, van Smeden M, De Cock B, Steyerberg EW. Regression shrinkage methods for clinical prediction models do not guarantee improved performance: Simulation study. Stat Methods Med Res. 2020;29(11):3166–78.

42. Collins GS, Ogundimu EO, Cook JA, Manach YL, Altman DG. Quantifying the impact of different approaches for handling continuous predictors on the performance of a prognostic model. Stat Med. 2016;35(23):4124–35.

43. Bennette C, Vickers A. Against quantiles: categorization of continuous variables in epidemiologic research, and its discontents. BMC Med Res Methodol. 2012;12:21.

44. Altman DG, Royston P. The cost of dichotomising continuous variables. BMJ. 2006;332(7549):1080.

45. Royston P, Altman DG, Sauerbrei W. Dichotomizing continuous predictors in multiple regression: a bad idea. Stat Med. 2006;25(1):127–41.

46. Sauerbrei W, Royston P, Binder H. Selection of important variables and determination of functional form for continuous predictors in multivariable model building. Stat Med. 2007;26(30):5512–28.

47. Whittle R, Royle K-L, Jordan KP, Riley RD, Mallen CD, Peat G. Prognosis research ideally should measure time-varying predictors at their intended moment of use. Diagnostic and Prognostic Research. 2017;1(1):1.

48. Yuan W, Beaulieu-Jones BK, Yu KH, Lipnick SL, Palmer N, Loscalzo J, et al. Temporal bias in case-control design: preventing reliable predictions of the future. Nat Commun. 2021;12(1):1107.

49. de Hond AAH, Shah VB, Kant IMJ, Van Calster B, Steyerberg EW, Hernandez-Boussard T. Perspectives on validation of clinical predictive algorithms. NPJ Digit Med. 2023;6(1):86.

50. Moons KG, Kengne AP, Grobbee DE, Royston P, Vergouwe Y, Altman DG, et al. Risk prediction models: II. External validation, model updating, and impact assessment. Heart. 2012;98(9):691–8.

51. Riley RD, Ensor J, Snell KI, Debray TP, Altman DG, Moons KG, et al. External validation of clinical prediction models using big datasets from e-health records or IPD meta-analysis: opportunities and challenges. BMJ. 2016;353:i3140.

52. Van Calster B, Steyerberg EW, Wynants L, van Smeden M. There is no such thing as a validated prediction model. BMC Med. 2023;21(1):70.

53. Collins GS, Ogundimu EO, Altman DG. Sample size considerations for the external validation of a multivariable prognostic model: a resampling study. Stat Med. 2016;35(2):214–26.

54. Riley RD, Snell KIE, Archer L, Ensor J, Debray TPA, van Calster B, et al. Evaluation of clinical prediction models (part 3): calculating the sample size required for an external validation study. BMJ. 2024;384:e074821.

55. Riley RD, Debray TPA, Collins GS, Archer L, Ensor J, van Smeden M, et al. Minimum sample size for external validation of a clinical prediction model with a binary outcome. Stat Med. 2021;40(19):4230–51.

56. Van Calster B, McLernon DJ, van Smeden M, Wynants L, Steyerberg EW, Topic Group ‘Evaluating diagnostic t, et al. Calibration: the Achilles heel of predictive analytics. BMC Med. 2019;17(1):230.

57. Van Calster B, Nieboer D, Vergouwe Y, De Cock B, Pencina MJ, Steyerberg EW. A calibration hierarchy for risk models was defined: from utopia to empirical data. J Clin Epidemiol. 2016;74:167–76.

58. Van Calster B, Vickers AJ. Calibration of risk prediction models: impact on decision-analytic performance. Med Decis Making. 2015;35(2):162–9.

59. Andaur Navarro CL, Damen JAA, van Smeden M, Takada T, Nijman SWJ, Dhiman P, et al. Systematic review identifies the design and methodological conduct of studies on machine learning-based prediction models. J Clin Epidemiol. 2023;154:8–22.

60. Collins GS, Moons KGM, Dhiman P, Riley RD, Beam AL, Van Calster B, et al. TRIPOD+AI statement: updated guidance for reporting clinical prediction models that use regression or machine learning methods. BMJ. 2024;385:e078378.

61. Collins GS, Dhiman P, Andaur Navarro CL, Ma J, Hooft L, Reitsma JB, et al. Protocol for development of a reporting guideline (TRIPOD-AI) and risk of bias tool (PROBAST-AI) for diagnostic and prognostic prediction model studies based on artificial intelligence. BMJ Open. 2021;11(7):e048008.

62. Varney S. HIV preventive care is supposed to be free in teh US. So, why are some patients still paying? KFF Health News [Internet]. 2022 August 8, 2024. Available from: https://kffhealthnews.org/news/article/prep-hiv-prevention-costs-covered-problems-insurance/.

63. Braidwood Management, Incorporated; John Scott Kelley; Kelley Orthodontics; Ashley Maxwell; Zach Maxwell; Joel Starnes; Joel Miller; Gregory Scheideman versus Xavier Becerra, Secretary, U.S. Department of Health and Human Services; Janet Yellen, Secretary, U.S. Department of Treasury; Julie A. Su, Acting Secretary, U.S. Department of Labor. United States Court of Appeals for the Fifth Circuit; 2024.

64. Wan X, Wang W, Liu J, Tong T. Estimating the sample mean and standard deviation from the sample size, median, range and/or interquartile range. BMC Med Res Methodol. 2014;14:135.

