## SUPPLEMENTARY MATERIALS for "Electronic Health Record-Based Prediction Models to Inform Decisions about HIV Pre-exposure Prophylaxis: A Systematic Review"

**APPENDIX**

**Appendix 1.** Search strategy in PubMed and CINAHL

**Appendix 2.** PICOTS (populations, interventions, comparison, outcomes, timing, and setting) criteria used in defining the scope of the review

**Appendix 3.** Risk of bias assessment (PROBAST Short Form) of electronic health record-based prediction models for HIV infection

**Appendix 1. Search strategy in PubMed and CINAHL**

1. **Database: PubMed** (Search date: June 19, 2023)

((HIV OR "Acquired Immunodeficiency Syndrome" OR "people living with HIV") AND (EHR OR EMR OR "electronic health records" OR "electronic medical records")) AND (Clinical Prediction Guides/Broad [filter])

2. **Database: CINAHL (**Search date: June 19, 2023)

| **#** | **Query** | **Results** |
| --- | --- | --- |
| 1 | (MH “Clinical Prediction Rules”) OR (MH “Decision Making, Clinical”) OR (MH “Decision Support Systems, Clinical”) | 41,674 |
| 2 | HIV OR Acquired Immunodeficiency syndrome OR People Living with HIV | 136,823 |
| 3 | EHR OR EMR OR Electronic Health Records OR Electronic Medical Record | 48,815 |
| 4 | (HIV OR Acquired Immunodeficiency syndrome OR People Living with HIV) AND (EHR OR EMR OR Electronic Health Records OR Electronic Medical Record) | 670 |
| 5 | ((MH “Clinical Prediction Rules”) OR (MH “Decision Making, Clinical”) OR (MH “Decision Support Systems, Clinical”)) AND ((HIV OR Acquired Immunodeficiency syndrome OR People Living with HIV) AND (EHR OR EMR OR Electronic Health Records OR Electronic Medical Record)) | 22 |

**Appendix 2. PICOTS (populations, interventions, comparison, outcomes, timing, and setting) criteria used in defining the scope of the review**

| **Population** | Any adults ( ≥18 years) without a prior diagnosis of HIV at baseline and are at high risk of HIV infection |
| --- | --- |
| **Intervention(s)** | Any development or validation of a prognostic risk prediction model that predicts the future risk of HIV infection (i.e., incidence) or is aimed at identifying individuals who are at high risk of HIV infection and may benefit from PrEP. |
| **Comparison (if applicable)** | Not applicable |
| **Outcome(s)** | HIV infection |
| **Timing** | Future risk of HIV infection (i.e., incidence) is predicted using predictors available to the clinician at the time of the care visit. |
| **Setting** | Any setting healthcare-related setting with EHR use |

**Appendix 3. Risk of bias assessment of electronic health record-based prediction models for HIV infection**

| **Domain** | | **Feller**  **(2018)** | **Krakower(2019)** | **Marcus**  **(2019)** | **Duthe**  **(2021)** | **Burns**  **(2022)** | **Friedman**  **(2023)** | **Xu**  **(2022)** |
| --- | --- | --- | --- | --- | --- | --- | --- | --- |
| **Outcome Assessment** | 3.1 Was the outcome determined appropriately? | 0 | 0 | 0 | NI | 0 | 0 | 0 |
|  | 3.2 Was a pre-specified or standard outcome definition used? | 0 | 0 | 0 | NI | 0 | 0 | 0 |
|  | 3.4 Was the outcome defined and determined in a similar way for all participants? | 0 | 0 | 0 | NI | 0 | 0 | 0 |
| **Analysis** | 4.1 Were there a reasonable number of participants with the outcome? | 1 | 1 | 1 | 1 | 1 | 1 | 1 |
|  | 4.2 Were continuous and categorical predictors handled appropriately | 1 | 0 | 1 | 1 | 1 | 1 | 1 |
|  | 4.4 Were participants with missing data handled appropriately? | 1 | 1 | 1 | 1 | 1 | 1 | 1 |
|  | 4.5 Was selection of predictors based on univariable analysis avoided?* | 0 | 0 | 0 | 0 | 0 | 1 | 0 |
|  | 4.8 Were model overfitting and optimism in model performance accounted for?* | 1 | 0 | 0 | 0 | 1 | 1 | 0 |
|  | Overall | High ROB | High  ROB | High ROB | High ROB | High ROB | High ROB | High ROB |

0 = No risk of bias

≥1= High risk of bias

All studies had small EPV so points were counted for item 4.5
